# Characterizing co-occurring conditions by age at diagnosis in autism spectrum disorders

**DOI:** 10.1101/19002527

**Authors:** Michelle D. Failla, Kyle L. Schwartz, Shikha Chaganti, Laurie E. Cutting, Bennett A. Landman, Carissa J. Cascio

**Affiliations:** Department of Psychiatry and Behavioral Science, Vanderbilt University Medical Center, Nashville, TN, USA; Departments of Electrical Engineering and Computer Science, Vanderbilt University, Nashville, TN; Department of Special Education, Vanderbilt University, Nashville, TN, USA; Vanderbilt Kennedy Center, Vanderbilt University, Nashville, TN, USA

## Abstract

Individuals with autism spectrum disorders (ASD) experience a significant number of co-occurring medical conditions, yet little is known about these conditions beyond prevalence. We hypothesized that individuals with ASD experienced an increased burden of co-occurring conditions as measured by presence, frequency, and duration of visits related to co-occurring conditions. We expected that age of ASD diagnosis (early, <7; late, >7) would be associated with different co-occurring conditions. Medical record data were extracted from a large anonymized medical center database for 3097 individuals with ASD and 3097 matched controls. Co-occurring conditions were characterized using a novel tool (pyPheWAS) to examine presence, frequency, and duration of each condition. We identified several categories of co-occurring conditions in ASD: neurological (epilepsy, sleep disorders); psychiatric (mood disorders, adjustment/conduct disorders, suicidal ideation), and developmental. Early ASD diagnosis was associated with epilepsy-related conditions, whereas a later diagnosis was associated with psychiatric conditions. The early ASD diagnosis group had later first diagnosis of co-occurring psychiatric conditions compared to the late ASD diagnosis group. Our work confirms individuals with ASD are under a significant medical burden, with increased duration and frequency of visits associated with co-occurring conditions. Adequate management of these conditions could reduce burden on individuals with ASD.

## Introduction

Autism spectrum disorders (ASD) are characterized by altered social communication as well as restrictive and repetitive behaviors. Patients with ASD often present with a wide and varied profile of co-occurring medical conditions^1–5^ that can greatly impact them and their families. Commonly reported co-occurring conditions in ASD include developmental delays^6^, gastrointestinal issues^7^, epilepsy^5^, and other psychiatric conditions^8^. Recent studies have used electronic medical records (EMR) to examine co-occurring conditions in ASD^2,9–11^, often corroborating previous findings from smaller cohorts^3,8,12^. In this study, we extend the characterization of co-occurring medical conditions in ASD, moving beyond prevalence to examining onset, duration, and frequency of visits associated with each co-occurring condition. Characterizing medical burden in this way may lead to better, more cost-effective care.

Individuals with ASD report being unsatisfied with their healthcare experience, especially with management of long-term conditions^13^. Management of co-occurring medical conditions with ASD increases healthcare costs and burdens on family and other care-takers^14^, and many families describe difficulty in accessing appropriate care^15^. Compared to adults without ASD, individuals with ASD are more likely to visit the emergency department^13^, most often due to co-occurring conditions^16^, and, on average, incur higher costs^16^. Thus, understanding how and when co-occurring medical conditions present, and how they are managed, could reduce cost and mitigate burden on individuals and families.

As patients with ASD are highly heterogeneous, subtyping can allow for earlier identification, better prognostication, and more personalized treatment. Previous work using EMR has characterized ASD subgroups by presence of co-occurring conditions^11,17^. We hypothesized that age at ASD diagnosis may differentiate profiles of co-occurring conditions. Later age of ASD diagnosis is associated with more co-occuring conditions^6,18^. While estimates of average age at ASD diagnosis range from 3.1-7.2 years^19^, there is a growing subset of individuals who may not receive an ASD diagnosis until adulthood^20,21^. It is not clear if individuals diagnosed in late childhood^22^, or even adulthood^23^, represent phenotypically distinct subtypes, missed/erroneous diagnoses, subtle presentation^22,24^, or some combination of other factors. Thus, we examined co-occurring conditions in those with an early (prior to age 7) and late ASD diagnosis (after age 7) to further characterize co-occurring conditions across different cohorts. We considered several factors in our diagnostic age split: the distribution of age at diagnosis in our data set, previous estimates of average age at diagnosis^19^, previous co-occurring condition studies^18^, and potential life-events (eg. starting school) that could influence identification.

In this study, we used a newly developed tool for co-occurring condition assessment in EMR^25^ to characterize medical burden by the presence of these conditions, as well as the frequency and duration of visits associated with each condition. We provide evidence for increased burden of co-occurring medical conditions in ASD, overall, and varying profiles of co-occurring condition burden based on age at ASD diagnosis.

## Methods

### Patient Sample

This study was approved by the Internal Review Board of Vanderbilt University Medical Center. We compared individuals with ASD (n=3097) to typically developing (TD) controls, matched on age, sex, and race (n=3097). **Figure 1** describes the study flow. The complete International Classification of Diseases, Ninth Revision (ICD-9) history was obtained for each patient from de-identified electronic health records at Vanderbilt University Medical Center (1991-2015). Electronic health records included in the Vanderbilt University Medical Center system include ∼2,000,000 visits per year in 120 inpatient and outpatient facilities across the Mid-South. Demographics are reported in **Table 1**. Individuals with ASD had at least one ICD-9 code for ASD (299, 299.0, 299.00, 299.01, 299.1, 299.10, 299.11, 299.8, 299.80, 299.81, 299.9, 299.90, 299.91), similar to previous work^11^. As some EMR studies in ASD have reported cohorts requiring at least two ICD-9 codes for the condition of interest^2,9,10^, we replicated the primary findings in this report in a smaller cohort that met this criterion (see Supplemental Table 1.)

**Table 1.** Demographic information for the ASD and control cohorts.

|  |  | Total |  |  | Lifespan |  |  | Early ASD Dx |  |  | Late ASD Dx |  |  |
| --- | --- | --- | --- | --- | --- | --- | --- | --- | --- | --- | --- | --- | --- |
|  |  | TD<br>n=2847 | ASD<br>n=2816 | *p* | TD<br>n=2210 | ASD<br>n=1474 | *p* | TD<br>n=1280 | ASD<br>n=831 | *p* | TD<br>n=909 | ASD<br>n=704 | *p* |
| **Sex** | Female | 566 | 555 | 0.987 | 417 | 267 | 0.844 | 209 | 134 | 0.460 | 188 | 173 | 0.169 |
|  | Male | 2276 | 2256 |  | 1790 | 1152 |  | 1071 | 696 |  | 719 | 530 |  |
|  | Unknown | 5 | 5 |  | 3 | 1 |  | 0 | 1 |  | 2 | 1 |  |
| **Race** | White | 1639 | 1430 | <0.001 | 1285 | 785 | <0.001 | 754 | 486 | 0.058 | 538 | 431 | 0.088 |
|  | Black/African American | 411 | 224 |  | 317 | 138 |  | 172 | 85 |  | 132 | 70 |  |
|  | Asian | 23 | 17 |  | 20 | 9 |  | 3 | 1 |  | 7 | 5 |  |
|  | Hispanic | 138 | 94 |  | 112 | 54 |  | 71 | 42 |  | 31 | 24 |  |
|  | American Indian or Alaska Native | 4 | 4 |  | 3 | 3 |  | 13 | 5 |  | 0 | 0 |  |
|  | Unknown | 632 | 1047 |  | 473 | 431 |  | 267 | 289 |  | 201 | 174 |  |
| **Death Status** | Alive | 2827 | 2802 | 0.408 | 2200 | 1471 | 0.367 | 1277 | 829 | 0.999 | 900 | 700 | 0.509 |
|  | Deceased | 20 | 14 |  | 10 | 3 |  | 3 | 2 |  | 9 | 4 |  |

**Figure 1.**
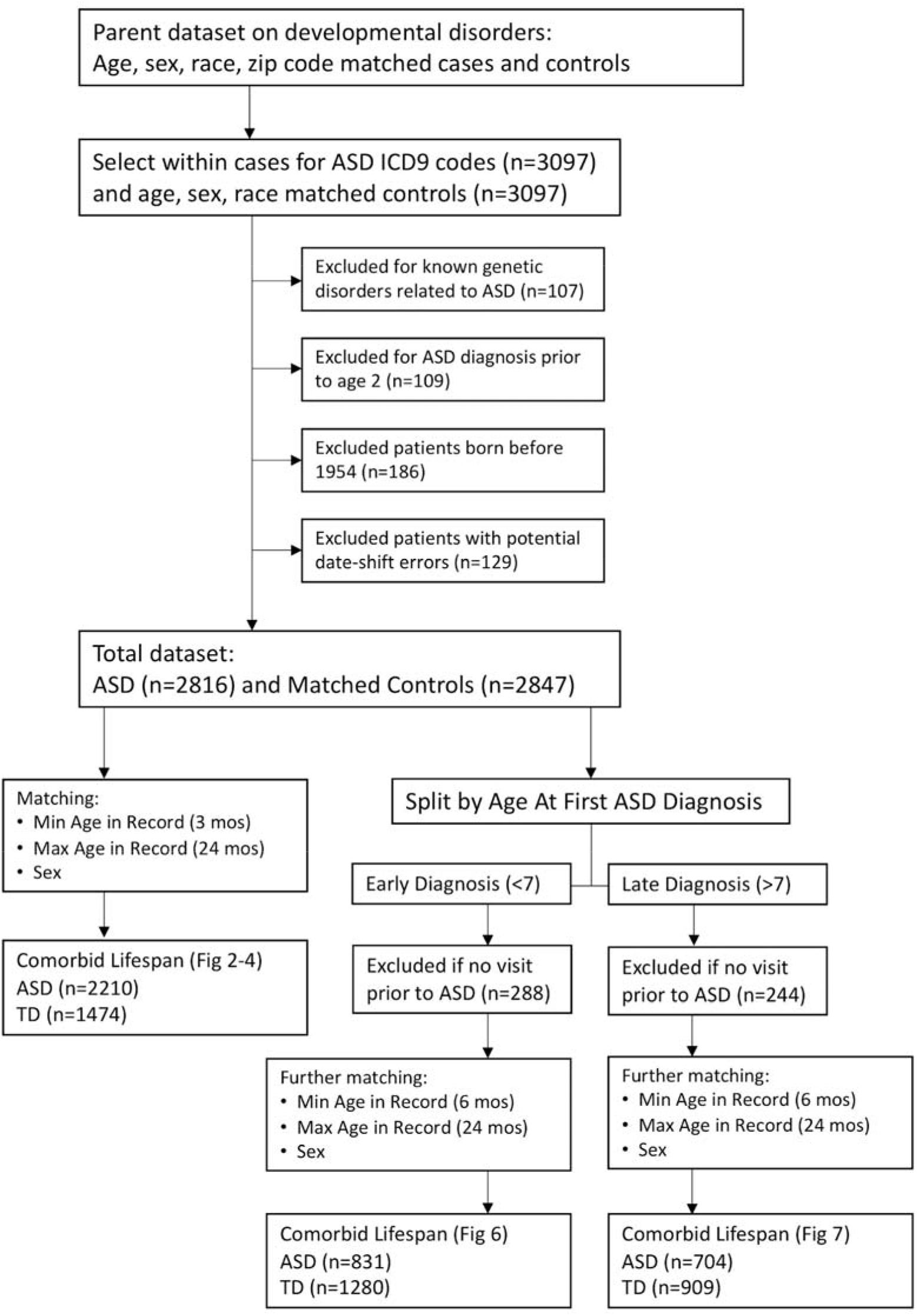
Overall study design.

### Exclusion criteria

Approximately 5% of individuals with ASD have a potentially causative genetic condition^26^ that could increase the likelihood of certain medical conditions. Thus, we excluded patients with Rett’s Syndrome (330.8), Chromosomal and congenital abnormalities (758.x, 759.89), Tuberous Sclerosis (759.5), and Fragile-X (759.83). We also excluded patients who received an ASD code prior to age 2 as diagnoses before age 2 are not generally stable^27^. We removed patients born before 1954 and records with potential date shift errors (usually related to short EMRs around the time of birth). **Figure 1** describes the number of patients excluded for each reason.

### PheDAS Analysis

EMR phenotypes called *phecodes*^17,28^ were identified from anonymized EMRs based on ICD-9 codes. EMR phenotypes are sets of characteristics/symptoms that consistently define clinical presentations of different conditions. Using a new python tool (pyPheWAS), significant conditions were identified using phenome-disease association study (PheDAS^25^), wherein associations between each EMR phenotype and the condition of interest (ASD) are tested. Individuals’ ICD-9 codes were mapped to phecodes and the incidence of each condition was determined relative to matched controls. Logistic regression, with ASD group membership as the dependent variable, was used to examine relative association of phecodes with the ASD cohort compared to the control cohort. We conducted three sets of logistic regressions where the phecode variable was treated in one of three ways: as presence (binary absence/presence of the code), count/frequency (number of incidences of the code), or duration (min-max dates the code appeared). For each set of regressions, PheDAS runs 1865 logistic regressions (one for each phecode) with each regression yielding a p-value indicating association between the condition and ASD. Multiple comparison correction was conducted with Bonferroni.

### Analysis by Age at Diagnosis

Age at ASD diagnosis was explored as a potential factor in co-occurring condition profiles. Age at ASD diagnosis was defined as the age at first ASD phecode in the EMR. The ASD cohort was split by age at diagnosis: first ASD code before age 7 (early diagnosis) and after age 7 (late diagnosis). For this analysis, we only included patients who had at least 1 visit prior to their first ASD code to increase confidence in the first ASD code as a diagnostic event. See **Table 1** for demographics for each cohort.

### Additional Statistical Analyses

To determine differences in demographic variables between ASD and controls and between early and late diagnostic groups, we used Chi-square tests. We further examined two categories of phecodes: epilepsy-related conditions (including any codes related to epilepsy, recurrent seizures, convulsions; generalized convulsive epilepsy; convulsions; epilepsy; and encephalopathy, not elsewhere classified) and psychiatric conditions (including any codes related to mood disorder; anxiety disorder; major depressive disorder; anxiety, phobic, and dissociative disorders; other mental disorders; adjustment disorders; conduct disorders; and suicidal ideation). We examined the proportion of individuals with each co-occurring condition type by diagnostic cohort using a Chi-square test. Mean onset times (first diagnostic code in our dataset) for each condition were compared by ASD diagnostic group using Mann-Whitney. We calculated time between onset of epilepsy or psychiatric conditions (age at first condition-related phecode - age at first ASD phecode). Positive values indicate onset of co-occurring condition after ASD diagnosis, while negative values indicate onset of co-occurring conditions prior to ASD diagnosis. Analyses were conducted using Python (v.2.7) and R (v.3.3.2).

## Results

### Burden of co-occurring medical conditions across the lifespan in ASD

We investigated the presence of co-occurring conditions in ASD across the lifespan. The ASD group was matched to the TD group on age at first visit (within 3 months), age at last visit (within 24 months), and sex (up to 3 matched controls; n, ASD=1420, TD=2210, Lifespan Cohort, **Table 1**). Phecodes enriched in the ASD group included neurological disorders like seizure disorders, sleep disturbances; psychiatric disorders including mood disorders, suicidal ideation, adjustment reaction, anxiety disorders, psychosis, speech and language disorders; developmental delays; and other ill-defined and unknown causes of morbidity and mortality.

Several conditions were less prevalent in the ASD group: contusions, congenital abnormalities of great vessels, nonspecific chest pain, acute upper respiratory and viral infections, and injuries (**Figure 2, Table 2**, see **Supplemental Table 1** for similar results in a cohort where two ASD ICD-9 codes were required). The ASD group had more visits for neurological conditions (sleep disorders, insomnia, and convulsions), psychiatric conditions (mood disorders, pervasive developmental disorders, anxiety, conduct disorders), developmental delays and disorders, delayed milestones, and lack of normal physiological development. The ASD group had fewer visits for contusions, injuries, and other tests (**Figure 3, Table 3**). Several phecodes had longer durations in the ASD group: neurological conditions (encephalopathy, sleep disorders, insomnia, convulsions, and lack of coordination), psychiatric conditions (mood disorders, pervasive developmental disorders, developmental delays, adjustment reaction, conduct disorders, anxiety disorders, attention deficit hyperactivity disorder, transient alteration of awareness, speech and language disorders), developmental delays (lack of normal physiological development, delayed milestones). The TD group had longer durations of acute respiratory infections, injuries, and other tests (**Figure 4, Table 3**).

**Table 2.**
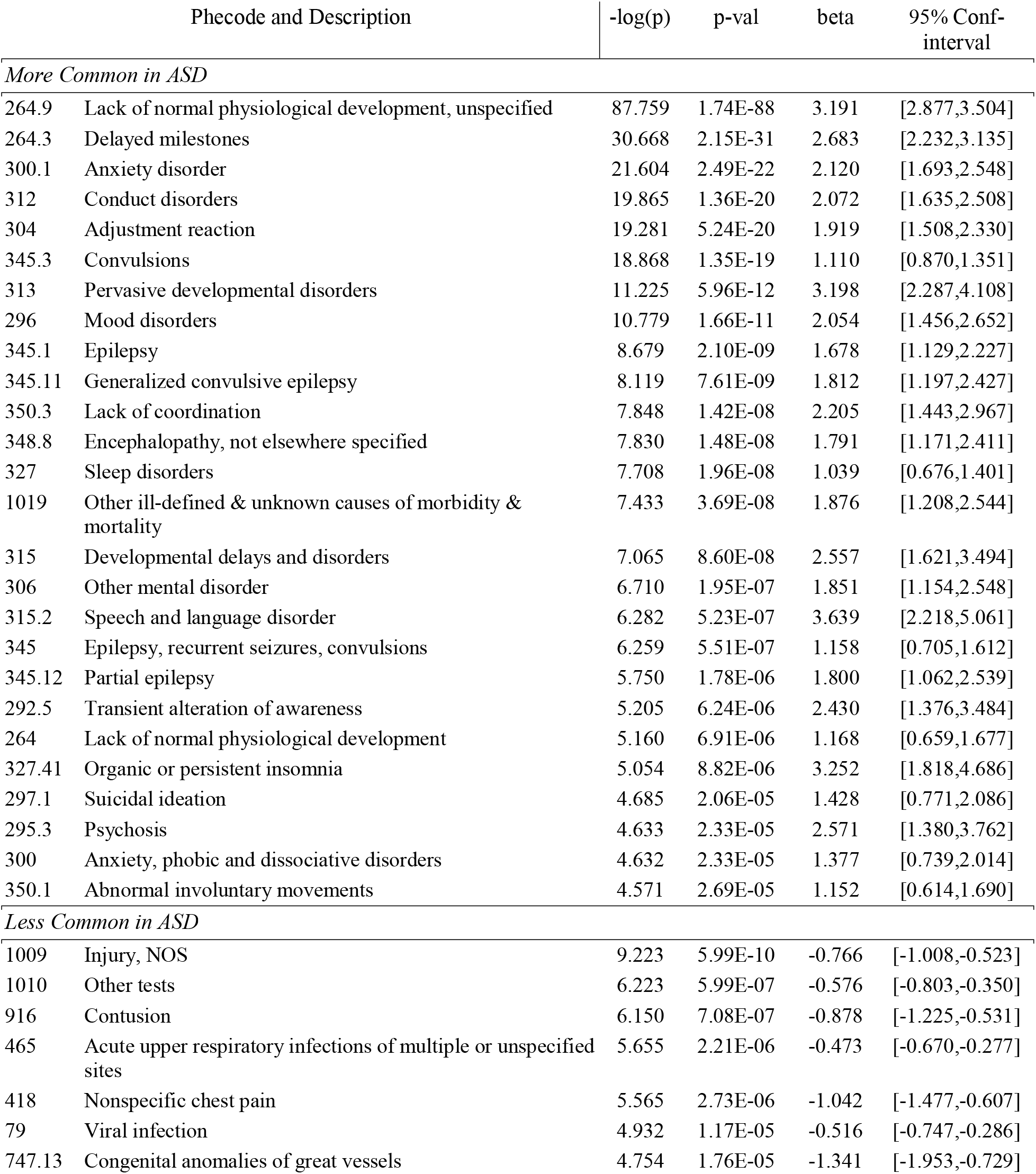
Regression models with the presence of co-occurring conditions associated with ASD.

**Table 3.** Regression models with the count of co-occurring conditions associated with ASD.

| Phecode and Description |  | -log(p) | p-val | beta | 95% Conf-interval |
| --- | --- | --- | --- | --- | --- |
| *Higher Count in ASD* |  |  |  |  |  |
| 264.9 | Lack of normal physiological development, unspecified | 69.455 | 3.51E-70 | 2.417 | [2.149,2.684] |
| 264.3 | Delayed milestones | 20.119 | 7.61E-21 | 1.512 | [1.196,1.829] |
| 312 | Conduct disorders | 12.837 | 1.46E-13 | 1.236 | [0.909,1.564] |
| 300.1 | Anxiety disorder | 10.887 | 1.30E-11 | 0.826 | [0.587,1.065] |
| 313 | Pervasive developmental disorders | 9.182 | 6.57E-10 | 2.284 | [1.559,3.009] |
| 345.3 | Convulsions | 7.517 | 3.04E-08 | 0.219 | [0.142,0.297] |
| 327 | Sleep disorders | 6.452 | 3.53E-07 | 0.612 | [0.377,0.848] |
| 315 | Developmental delays and disorders | 6.451 | 3.54E-07 | 2.434 | [1.497,3.371] |
| 306 | Other mental disorder | 6.140 | 7.24E-07 | 1.650 | [0.997,2.303] |
| 315.2 | Speech and language disorder | 5.506 | 3.12E-06 | 3.377 | [1.958,4.799] |
| 1019 | Other ill-defined and unknown causes of morbidity and mortality | 5.426 | 3.75E-06 | 1.271 | [0.732,1.810] |
| 296 | Mood disorders | 5.057 | 8.78E-06 | 0.289 | [0.161,0.416] |
| 327.41 | Organic or persistent insomnia | 4.604 | 2.49E-05 | 2.244 | [1.200,3.287] |
| *Lower Count in ASD* |  |  |  |  |  |
| 1010 | Other tests | 5.896 | 1.27E-06 | -0.162 | [-0.228,-0.097] |
| 1009 | Injury, NOS | 4.896 | 1.27E-05 | -0.232 | [-0.337,-0.128] |
| 916 | Contusion | 4.854 | 1.40E-05 | -0.563 | [-0.817,-0.309] |

**Table 4.**
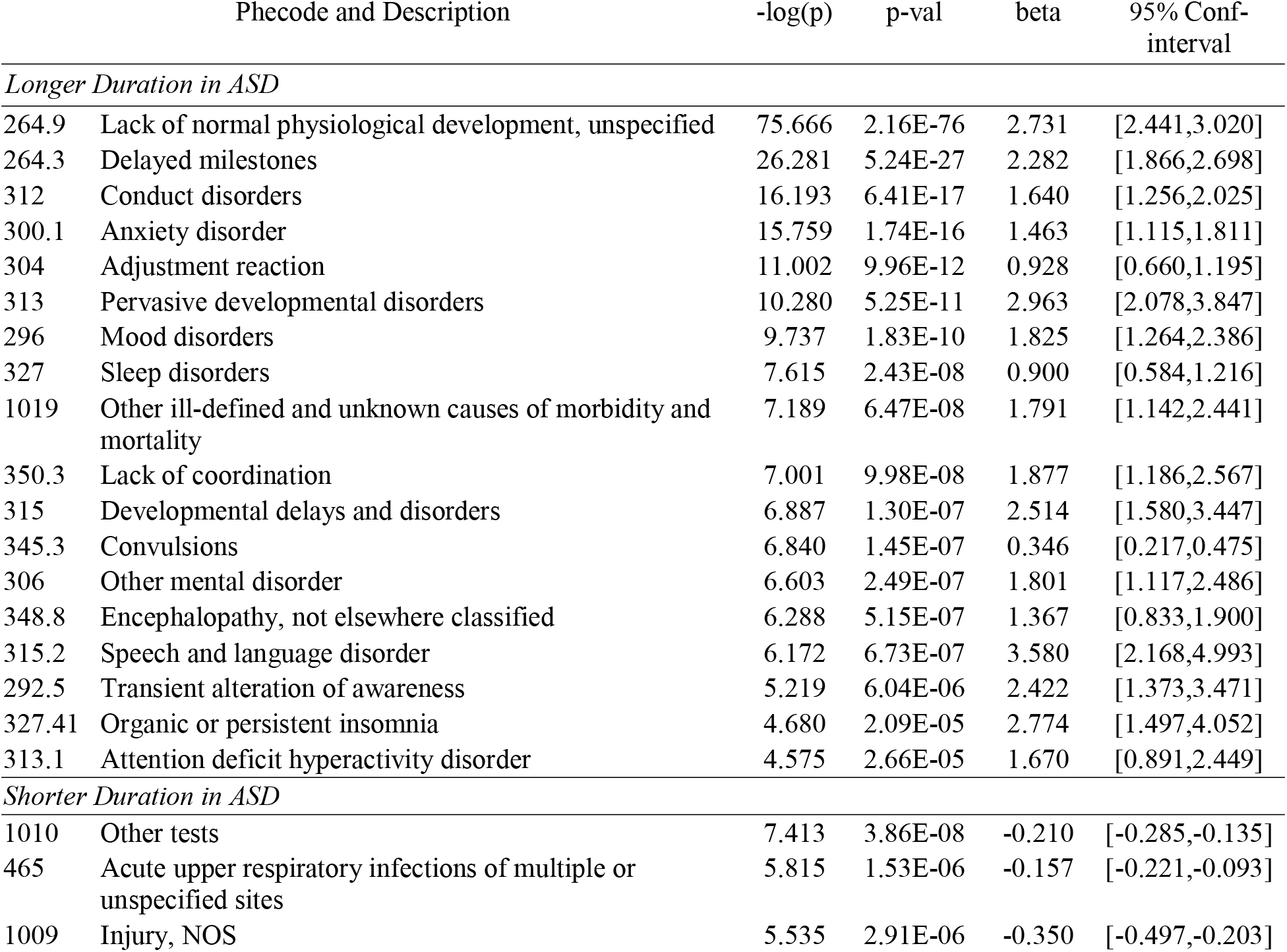
Regression models with the duration of co-occurring conditions associated with ASD.

| Phecode and Description |  | -log(p) | p-val | beta | 95% Conf-interval |
| --- | --- | --- | --- | --- | --- |
| *Longer Duration in ASD* |  |  |  |  |  |
| 264.9 | Lack of normal physiological development, unspecified | 75.666 | 2.16E-76 | 2.731 | [2.441,3.020] |
| 264.3 | Delayed milestones | 26.281 | 5.24E-27 | 2.282 | [1.866,2.698] |
| 312 | Conduct disorders | 16.193 | 6.41E-17 | 1.640 | [1.256,2.025] |
| 300.1 | Anxiety disorder | 15.759 | 1.74E-16 | 1.463 | [1.115,1.811] |
| 304 | Adjustment reaction | 11.002 | 9.96E-12 | 0.928 | [0.660,1.195] |
| 313 | Pervasive developmental disorders | 10.280 | 5.25E-11 | 2.963 | [2.078,3.847] |
| 296 | Mood disorders | 9.737 | 1.83E-10 | 1.825 | [1.264,2.386] |
| 327 | Sleep disorders | 7.615 | 2.43E-08 | 0.900 | [0.584,1.216] |
| 1019 | Other ill-defined and unknown causes of morbidity and mortality | 7.189 | 6.47E-08 | 1.791 | [1.142,2.441] |
| 350.3 | Lack of coordination | 7.001 | 9.98E-08 | 1.877 | [1.186,2.567] |
| 315 | Developmental delays and disorders | 6.887 | 1.30E-07 | 2.514 | [1.580,3.447] |
| 345.3 | Convulsions | 6.840 | 1.45E-07 | 0.346 | [0.217,0.475] |
| 306 | Other mental disorder | 6.603 | 2.49E-07 | 1.801 | [1.117,2.486] |
| 348.8 | Encephalopathy, not elsewhere classified | 6.288 | 5.15E-07 | 1.367 | [0.833,1.900] |
| 315.2 | Speech and language disorder | 6.172 | 6.73E-07 | 3.580 | [2.168,4.993] |
| 292.5 | Transient alteration of awareness | 5.219 | 6.04E-06 | 2.422 | [1.373,3.471] |
| 327.41 | Organic or persistent insomnia | 4.680 | 2.09E-05 | 2.774 | [1.497,4.052] |
| 313.1 | Attention deficit hyperactivity disorder | 4.575 | 2.66E-05 | 1.670 | [0.891,2.449] |
| *Shorter Duration in ASD* |  |  |  |  |  |
| 1010 | Other tests | 7.413 | 3.86E-08 | -0.210 | [-0.285,-0.135] |
| 465 | Acute upper respiratory infections of multiple or unspecified sites | 5.815 | 1.53E-06 | -0.157 | [-0.221,-0.093] |
| 1009 | Injury, NOS | 5.535 | 2.91E-06 | -0.350 | [-0.497,-0.203] |

**Figure 2.**
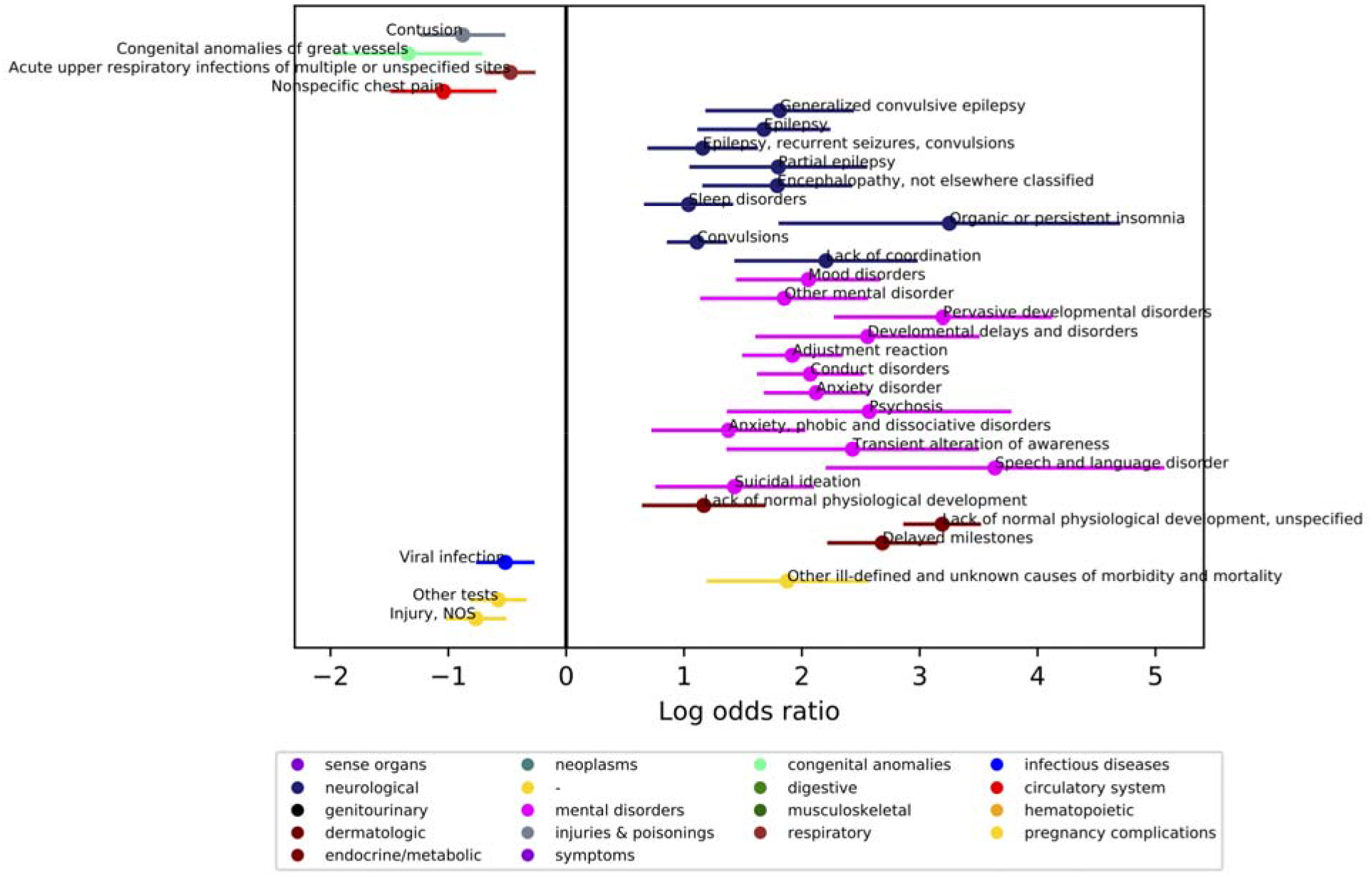
Comparing presence of medical conditions co-occurring with ASD diagnosis across the lifespan. Individuals with ASD were matched on age at first visit (within 3 months), age at last visit (within 24 months), and sex to a typical comparison group, up to 3 individuals in the TC group for each individual in the ASD group. (N, ASD=1420, Typical Comparison=2210). Phecodes with significant log odds ratios following a Bonferroni correction are displayed on the graph. Age at first visit was 4.51±6.70 years (range 0-48.22 years) and age at last visit was 8.85±7.28 years (range 0.11-59.72 years).

**Figure 3.**
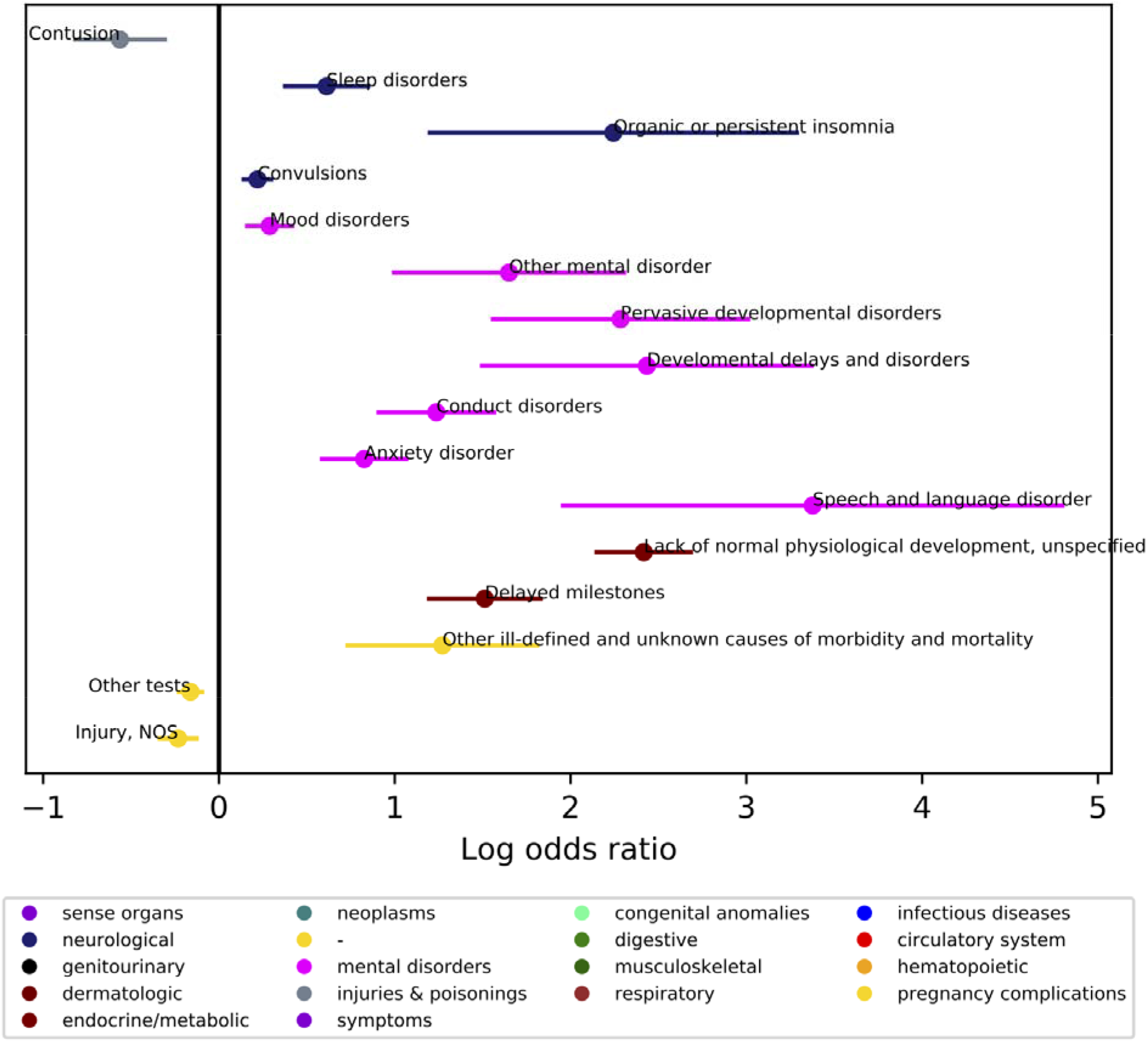
Comparing count incidences of medical conditions co-occurring with ASD diagnosis across the lifespan. Individuals with ASD were matched on age at first visit (within 3 months), age at last visit (within 24 months), and sex to a typical comparison group, up to 3 individuals in the TC group for each individual in the ASD group. (N, ASD=1420, Typical Comparison=2210). Phecodes with significant log odds ratios following a Bonferroni correction are displayed on the graph. Age at first visit was 4.51±6.70 years (range 0-48.22 years) and age at last visit was 8.85±7.28 years (range 0.11-59.72 years).

**Figure 4.**
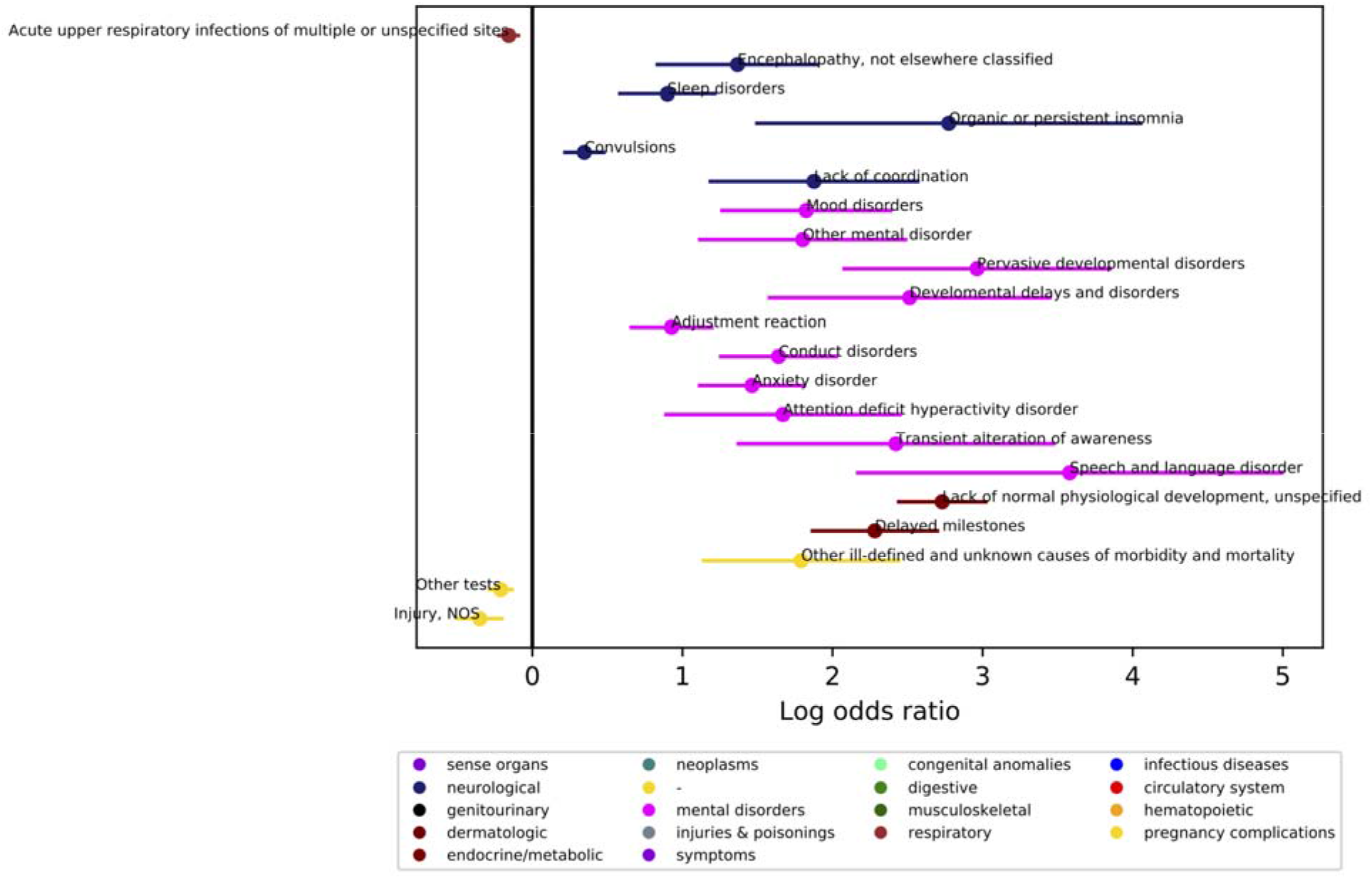
Comparing duration (minimum age the code first appears to maximum age the code last appears in the medical record) of medical conditions co-occurring with ASD diagnosis across the lifespan. Individuals with ASD were matched on age at first visit (within 3 months), age at last visit (within 24 months), and sex to a typical comparison group, up to 3 individuals in the TC group for each individual in the ASD group. (N, ASD=1420, Typical Comparison=2210). Phecodes with significant log odds ratios following a Bonferroni correction are displayed on the graph. Age at first visit was 4.51±6.70 years (range 0-48.22 years) and age at last visit was 8.85±7.28 years (range 0.11-59.72 years).

### Co-occurring medical condition profiles differ by age at diagnosis

The ASD group was split by age at diagnosis (<7, early; >7, late) and then each group was matched to TD on age at first visit (within 6 mos), age at last visit, (within 24 months), and sex. **Figure 5** shows the distribution of age at diagnosis by early (n=831) and late (n=704) ASD group. There were more females in the late diagnosis group (24.6% compared to 16.1%, □^2^ =17.04, p=0.0002; see **Table 1** for full demographics).

**Figure 5.**
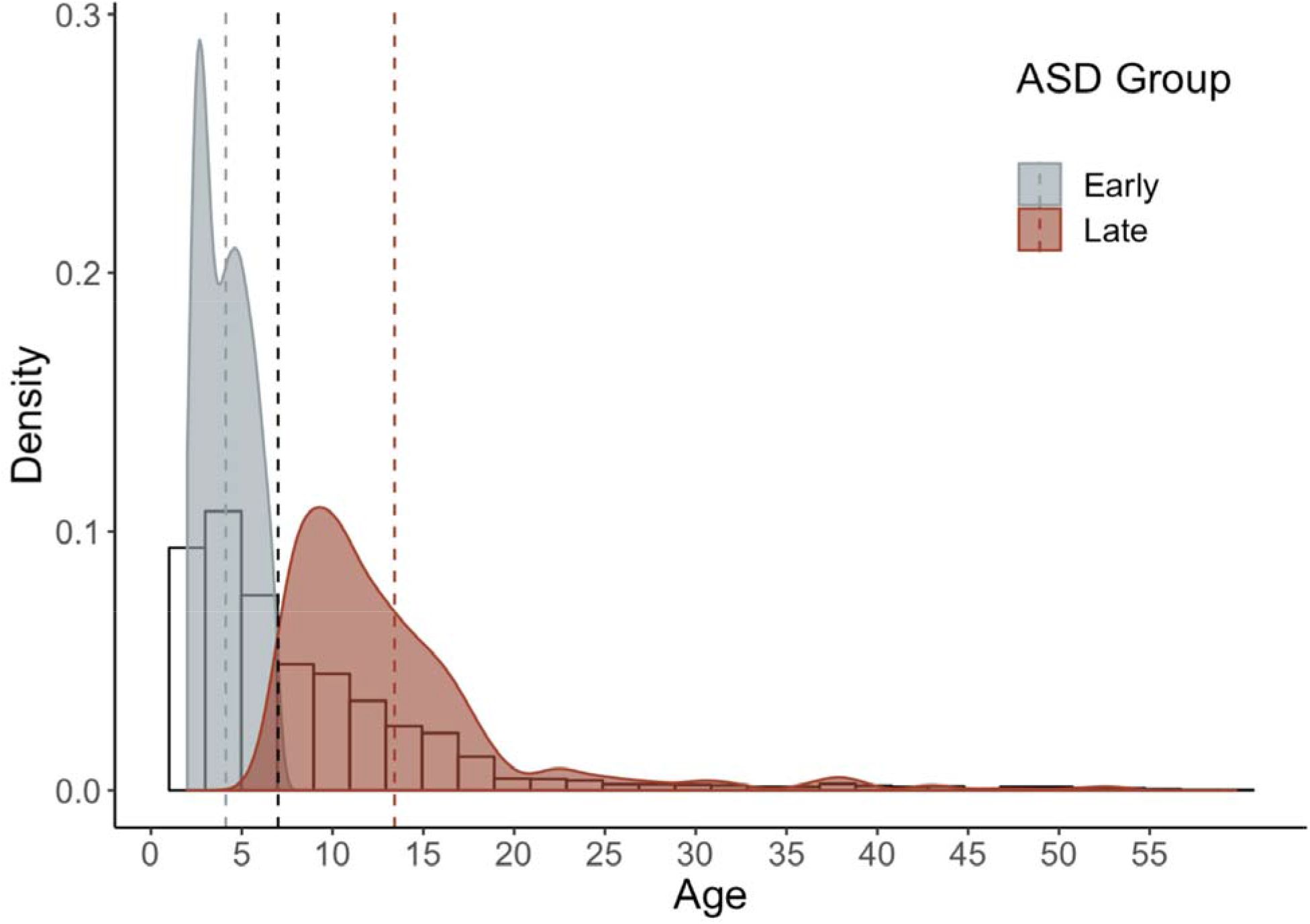
Histogram depicting age at first ASD diagnostic code. Dashed black line represents split at age of first diagnosis, before and after age 7. Histogram represents age at ASD diagnosis for total cohort (n=2847). Density plots indicate age at ASD diagnosis by diagnosis group, early: n=831, late: n=704; dashed lines designate mean age at diagnosis for each group.

We examined co-occurring conditions in the early diagnosis ASD group (**Figure 6, Table 5**). Phecodes enriched in the early ASD group included hearing disorders, neurological disorders: seizure disorders, encephalopathy, sleep disorders, lack of coordination; psychiatric disorders: pervasive developmental disorder, developmental delays and disorders, conduct disorders, anxiety disorder; and developmental codes: lack of normal physiological development, and delayed milestones. The early ASD group also had more visits for convulsions, pervasive developmental disorders, conduct disorders, developmental delays, lack of normal physiological development (unspecified), and delayed milestones. The early ASD group had longer durations for encephalopathy, sleep disorders, convulsions, pervasive developmental disorders, developmental delays and disorders, conduct disorders, anxiety disorders, lack of normal physiological development, and delayed milestones.

**Table 5.** Early ASD Diagnosis (< Age 7): Regression models of co-occurring conditions.

|  | Phecode and Description | -log(p) | p-val | beta | 95% Conf-interval |
| --- | --- | --- | --- | --- | --- |
| *More Common in ASD* |  |  |  |  |  |
| 264.9 | Lack of normal physiological development, unspecified | 76.939 | 1.15E-77 | 3.470 | [3.105,3.834] |
| 264.3 | Delayed milestones | 27.001 | 9.99E-28 | 2.656 | [2.179,3.133] |
| 345.3 | Convulsions | 13.753 | 1.77E-14 | 1.211 | [0.901,1.520] |
| 312 | Conduct disorders | 10.086 | 8.21E-11 | 1.802 | [1.258,2.345] |
| 313 | Pervasive developmental disorders | 9.924 | 1.19E-10 | 3.330 | [2.317,4.343] |
| 315 | Developmental delays and disorders | 7.413 | 3.87E-08 | 3.285 | [2.113,4.456] |
| 348.8 | Encephalopathy, not elsewhere classified | 6.559 | 2.76E-07 | 2.138 | [1.322,2.953] |
| 350.3 | Lack of coordination | 5.924 | 1.19E-06 | 2.354 | [1.404,3.304] |
| 345.1 | Epilepsy | 5.778 | 1.67E-06 | 1.741 | [1.029,2.453] |
| 389 | Hearing loss | 5.667 | 2.15E-06 | 1.044 | [0.612,1.475] |
| 264 | Lack of normal physiological development | 5.411 | 3.89E-06 | 1.528 | [0.879,2.177] |
| 327 | Sleep disorders | 5.363 | 4.33E-06 | 1.032 | [0.592,1.472] |
| 300.1 | Anxiety disorder | 5.243 | 5.72E-06 | 1.719 | [0.976,2.462] |
| 345.11 | Generalized convulsive epilepsy | 5.219 | 6.03E-06 | 1.919 | [1.088,2.750] |
| 1019 | Other ill-defined and unknown causes of morbidity and mortality | 4.924 | 1.19E-05 | 1.883 | [1.040,2.726] |
| *Higher Counts in ASD* |  |  |  |  |  |
| 264.9 | Lack of normal physiological development, unspecified | 61.582 | 2.62E-62 | 2.667 | [2.353,2.981] |
| 264.3 | Delayed milestones | 17.302 | 4.99E-18 | 1.412 | [1.093,1.732] |
| 313 | Pervasive developmental disorders | 8.686 | 2.06E-09 | 2.357 | [1.586,3.127] |
| 315 | Developmental delays and disorders | 6.914 | 1.22E-07 | 3.159 | [1.988,4.329] |
| 312 | Conduct disorders | 6.368 | 4.28E-07 | 0.998 | [0.611,1.385] |
| 345.3 | Convulsions | 4.654 | 2.22E-05 | 0.220 | [0.118,0.321] |
| *Longer Duration in ASD* |  |  |  |  |  |
| 264.9 | Lack of normal physiological development, unspecified | 66.840 | 1.45E-67 | 3.058 | [2.713,3.404] |
| 264.3 | Delayed milestones | 23.214 | 6.10E-24 | 2.247 | [1.811,2.684] |
| 345.3 | Convulsions | 9.217 | 6.07E-10 | 0.691 | [0.472,0.910] |
| 313 | Pervasive developmental disorders | 9.073 | 8.46E-10 | 3.077 | [2.094,4.060] |
| 315 | Developmental delays and disorders | 7.328 | 4.70E-08 | 3.248 | [2.082,4.413] |
| 312 | Conduct disorders | 7.183 | 6.56E-08 | 1.262 | [0.804,1.720] |
| 350.3 | Lack of coordination | 5.395 | 4.03E-06 | 2.065 | [1.187,2.943] |
| 327 | Sleep disorders | 5.261 | 5.48E-06 | 0.899 | [0.511,1.286] |
| 1019 | Other ill-defined and unknown causes of morbidity and mortality | 4.806 | 1.56E-05 | 1.767 | [0.965,2.568] |
| 300.1 | Anxiety disorder | 4.803 | 1.57E-05 | 1.587 | [0.867,2.308] |
| 348.8 | Encephalopathy, not elsewhere classified | 4.762 | 1.73E-05 | 1.508 | [0.820,2.195] |

**Figure 6.**
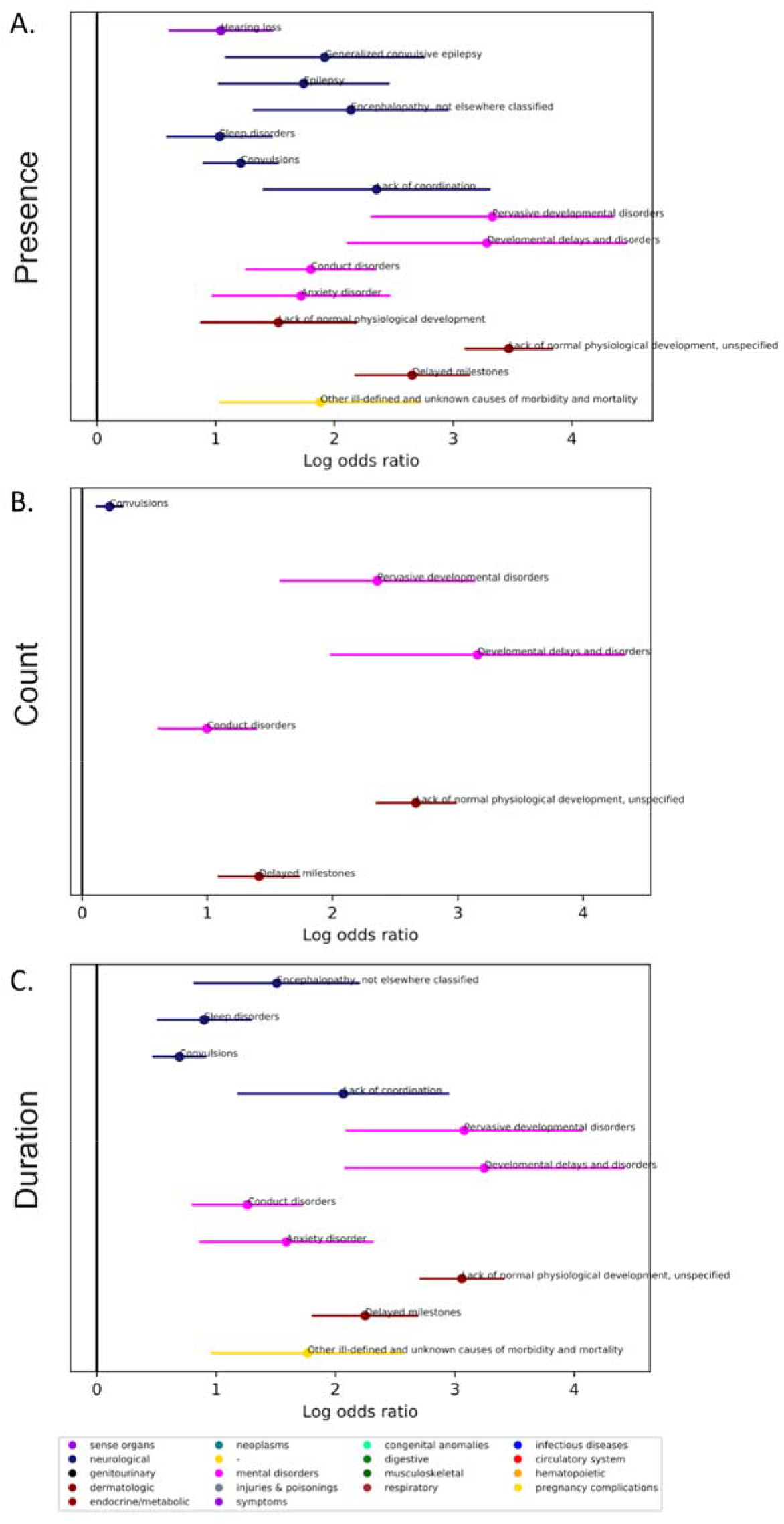
Comparing co-occurring medical conditions associated with ASD for those who were first diagnosed below age 7. Individuals with ASD were matched on age at first visit (within 6 months), age at last visit (within 24 months), and sex to a typical comparison (TC) group, up to two individuals in the TC group for each individual in the ASD group (N, ASD=831, Typical Comparison=1280). For the early diagnosis group, average age at first visit was 1.91±1.82 years (range 0-6.82 years) and average age at last visit was 5.70±2.72 years (range 2-18.48 years). Phecodes with significant log odds ratios following a Bonferroni correction are displayed on the graph. Phecodes are compared based on the presence in the record (top panel), the number of times the Phecode appears in the record (count, middle panel), and the length of time the Phecode is present in the record (duration, bottom panel).

We examined co-occurring conditions in the late diagnosed ASD group (**Figure 7, Table 6**). Phecodes enriched in the late ASD group included seizure disorders; psychiatric disorders: pervasive developmental disorder, mood disorders, depression, anxiety, adjustment reaction, conduct disorders, suicidal ideation; and developmental phecodes: lack of normal physiological development, and delayed milestones. The late ASD group had more visits related to convulsions, mood disorders, conduct disorders, anxiety disorders, lack of normal physiological development, and delayed milestones. The late ASD group also had longer durations for convulsions, mood disorders, adjustment disorders, conduct disorders, anxiety disorders, phobic and dissociative disorders, suicidal ideation, lack of normal physiological development, and delayed milestones. The late ASD group was less likely to have phecodes related to contusions (presence) and nonspecific injuries (presence and count).

**Table 6.**
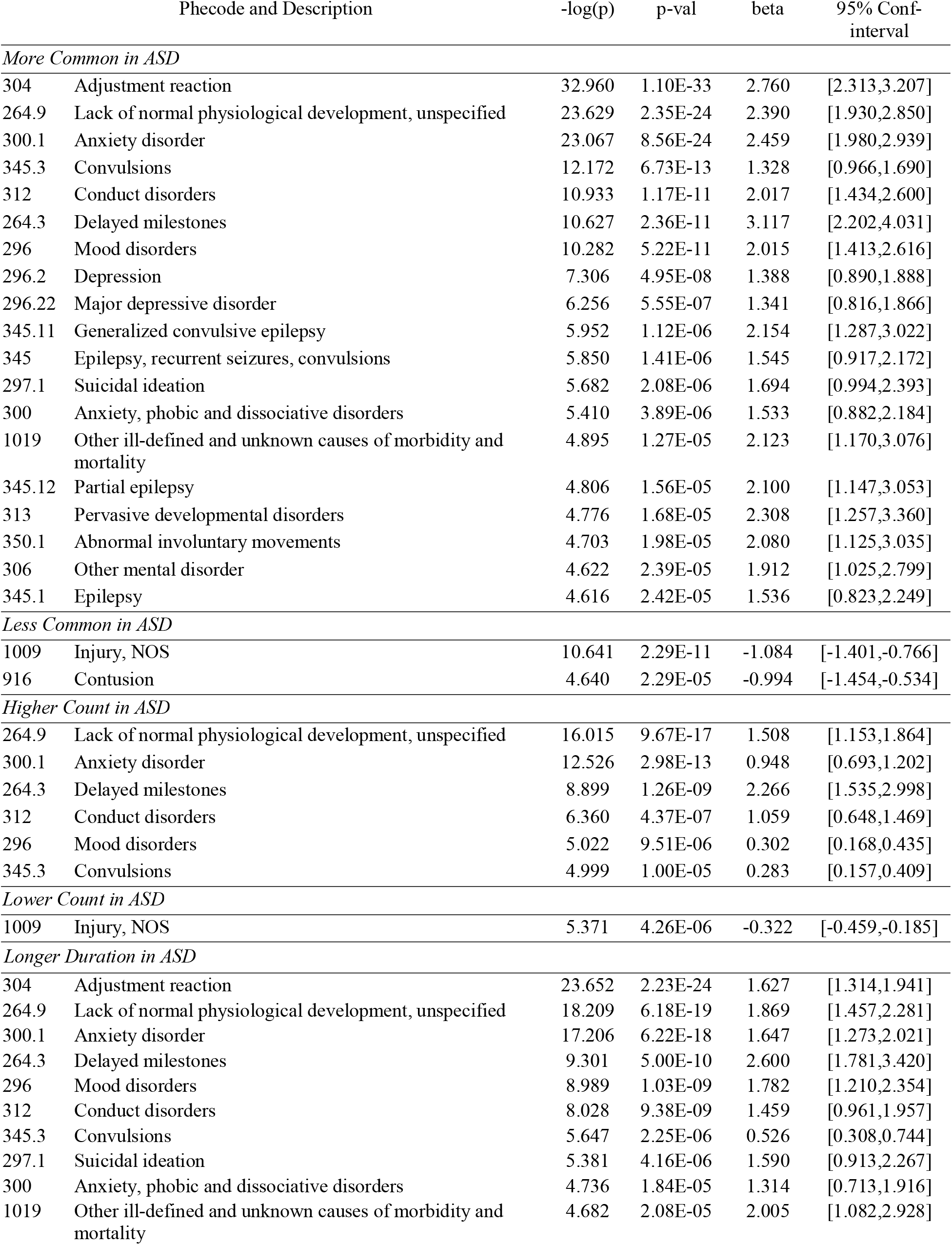
Late ASD Diagnosis (> Age 7): Regression models of co-occurring conditions.

| Phecode and Description |  | -log(p) | p-val | beta | 95% Conf-interval |
| --- | --- | --- | --- | --- | --- |
| *More Common in ASD* |  |  |  |  |  |
| 304 | Adjustment reaction | 32.960 | 1.10E-33 | 2.760 | [2.313,3.207] |
| 264.9 | Lack of normal physiological development, unspecified | 23.629 | 2.35E-24 | 2.390 | [1.930,2.850] |
| 300.1 | Anxiety disorder | 23.067 | 8.56E-24 | 2.459 | [1.980,2.939] |
| 345.3 | Convulsions | 12.172 | 6.73E-13 | 1.328 | [0.966,1.690] |
| 312 | Conduct disorders | 10.933 | 1.17E-11 | 2.017 | [1.434,2.600] |
| 264.3 | Delayed milestones | 10.627 | 2.36E-11 | 3.117 | [2.202,4.031] |
| 296 | Mood disorders | 10.282 | 5.22E-11 | 2.015 | [1.413,2.616] |
| 296.2 | Depression | 7.306 | 4.95E-08 | 1.388 | [0.890,1.888] |
| 296.22 | Major depressive disorder | 6.256 | 5.55E-07 | 1.341 | [0.816,1.866] |
| 345.11 | Generalized convulsive epilepsy | 5.952 | 1.12E-06 | 2.154 | [1.287,3.022] |
| 345 | Epilepsy, recurrent seizures, convulsions | 5.850 | 1.41E-06 | 1.545 | [0.917,2.172] |
| 297.1 | Suicidal ideation | 5.682 | 2.08E-06 | 1.694 | [0.994,2.393] |
| 300 | Anxiety, phobic and dissociative disorders | 5.410 | 3.89E-06 | 1.533 | [0.882,2.184] |
| 1019 | Other ill-defined and unknown causes of morbidity and mortality | 4.895 | 1.27E-05 | 2.123 | [1.170,3.076] |
| 345.12 | Partial epilepsy | 4.806 | 1.56E-05 | 2.100 | [1.147,3.053] |
| 313 | Pervasive developmental disorders | 4.776 | 1.68E-05 | 2.308 | [1.257,3.360] |
| 350.1 | Abnormal involuntary movements | 4.703 | 1.98E-05 | 2.080 | [1.125,3.035] |
| 306 | Other mental disorder | 4.622 | 2.39E-05 | 1.912 | [1.025,2.799] |
| 345.1 | Epilepsy | 4.616 | 2.42E-05 | 1.536 | [0.823,2.249] |
| *Less Common in ASD* |  |  |  |  |  |
| 1009 | Injury, NOS | 10.641 | 2.29E-11 | -1.084 | [-1.401,-0.766] |
| 916 | Contusion | 4.640 | 2.29E-05 | -0.994 | [-1.454,-0.534] |
| *Higher Count in ASD* |  |  |  |  |  |
| 264.9 | Lack of normal physiological development, unspecified | 16.015 | 9.67E-17 | 1.508 | [1.153,1.864] |
| 300.1 | Anxiety disorder | 12.526 | 2.98E-13 | 0.948 | [0.693,1.202] |
| 264.3 | Delayed milestones | 8.899 | 1.26E-09 | 2.266 | [1.535,2.998] |
| 312 | Conduct disorders | 6.360 | 4.37E-07 | 1.059 | [0.648,1.469] |
| 296 | Mood disorders | 5.022 | 9.51E-06 | 0.302 | [0.168,0.435] |
| 345.3 | Convulsions | 4.999 | 1.00E-05 | 0.283 | [0.157,0.409] |
| *Lower Count in ASD* |  |  |  |  |  |
| 1009 | Injury, NOS | 5.371 | 4.26E-06 | -0.322 | [-0.459,-0.185] |
| *Longer Duration in ASD* |  |  |  |  |  |
| 304 | Adjustment reaction | 23.652 | 2.23E-24 | 1.627 | [1.314,1.941] |
| 264.9 | Lack of normal physiological development, unspecified | 18.209 | 6.18E-19 | 1.869 | [1.457,2.281] |
| 300.1 | Anxiety disorder | 17.206 | 6.22E-18 | 1.647 | [1.273,2.021] |
| 264.3 | Delayed milestones | 9.301 | 5.00E-10 | 2.600 | [1.781,3.420] |
| 296 | Mood disorders | 8.989 | 1.03E-09 | 1.782 | [1.210,2.354] |
| 312 | Conduct disorders | 8.028 | 9.38E-09 | 1.459 | [0.961,1.957] |
| 345.3 | Convulsions | 5.647 | 2.25E-06 | 0.526 | [0.308,0.744] |
| 297.1 | Suicidal ideation | 5.381 | 4.16E-06 | 1.590 | [0.913,2.267] |
| 300 | Anxiety, phobic and dissociative disorders | 4.736 | 1.84E-05 | 1.314 | [0.713,1.916] |
| 1019 | Other ill-defined and unknown causes of morbidity and mortality | 4.682 | 2.08E-05 | 2.005 | [1.082,2.928] |

**Figure 7.**
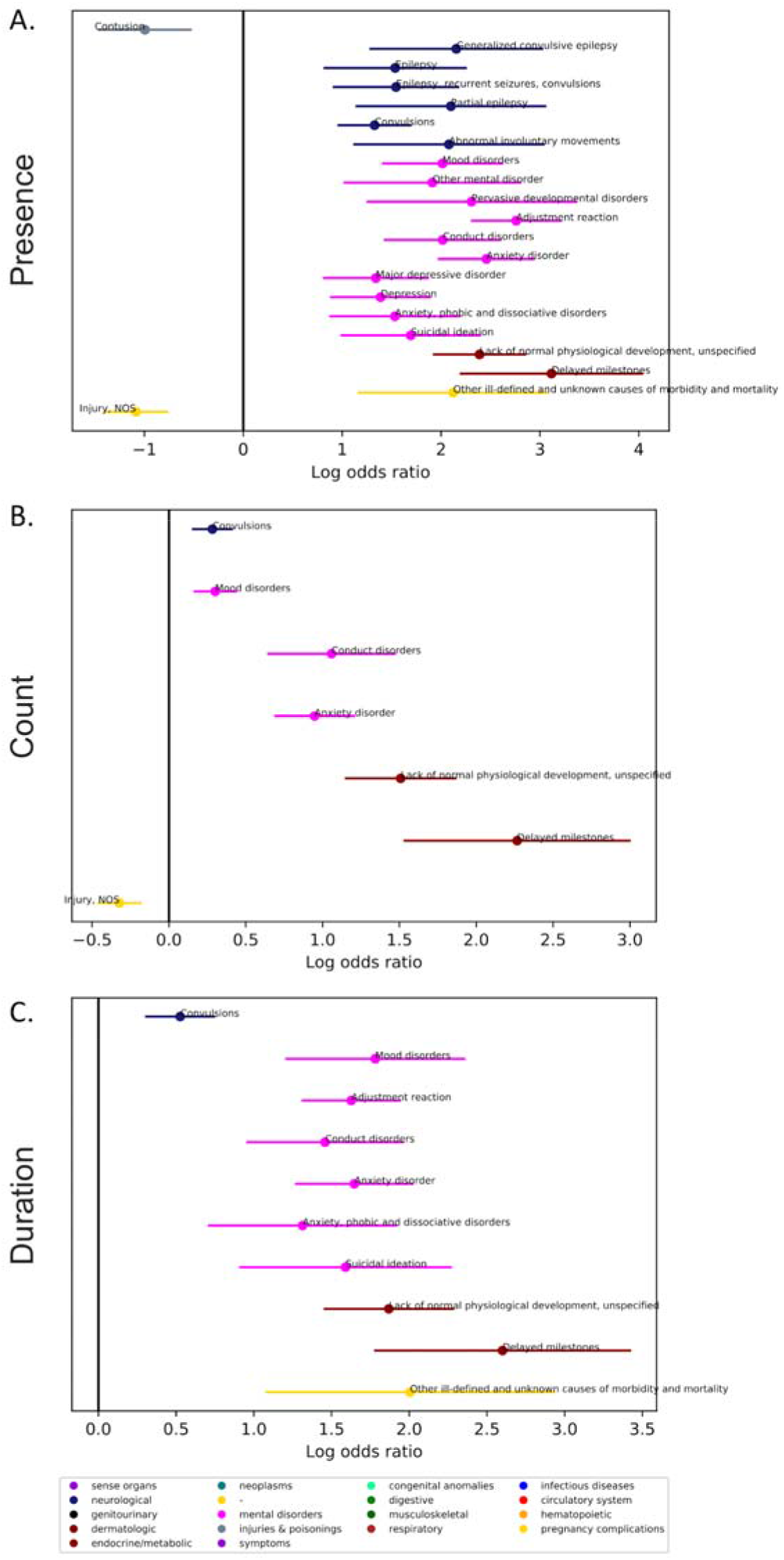
Comparing co-occurring medical conditions associated with ASD for those who were first diagnosed after age 7. Individuals with ASD were matched on age at first visit (within 6 months), age at last visit (within 24 months), and sex to a typical comparison group, up to two individuals in the TC group for each individual in the ASD group. (N, ASD=704, Typical Comparison=909). For the late diagnosis group, average age at first visit was 9.02±7.95 years (range 0-48.22 years) and average age at last visit was 14.93±7.67 years (range 7.01-59.04 years). Phecodes with significant log odds ratios following a Bonferroni correction are displayed on the graph. Phecodes are compared based on the presence in the record (top panel), the number of times the phecode appears in the record (count, middle panel), and the length of time the phecode is present in the record (duration, bottom panel).

### Further Characterization of Epilepsy-related and Psychiatric Conditions

We investigated differences related to two co-occurring condition categories: epilepsy-related conditions and psychiatric conditions. For epilepsy related codes, there was no significant difference in prevalence by age at ASD diagnosis (early diagnosis, 17.5%, compared to 18.5% for late diagnosis group, □^2^=0.15145, p=0.697). For psychiatric conditions, the late diagnosis group had significantly more psychiatric conditions (54.4%) compared to the early group (14.7%, □ ^2^=270.61, p=2.2e-16). We also examined first diagnostic code, or onset, of these conditions in relation to ASD diagnosis (**Figure 8**). For both diagnostic groups, epilepsy-related codes first appeared, on average, before the ASD diagnosis, (early: mean= -0.800 years, (1 standard deviation below/above mean, -2.955-1.355); late: mean= -2.271 years (1 standard deviation below/above mean, -6.646-2.104)), but earlier for the late ASD group (W=7941, p=0.019). There was a significant difference in the onset of psychiatric conditions by ASD group (W=16408, p=6.723e-07): in the early ASD group, psychiatric conditions first appeared, on average, *after* the diagnosis (mean=0.916 years, (−1.655-3.486)) whereas these conditions first appeared *before* the ASD diagnosis in the late ASD group (mean= -0.378 years, (−2.005-1.248)).

**Figure 8.**
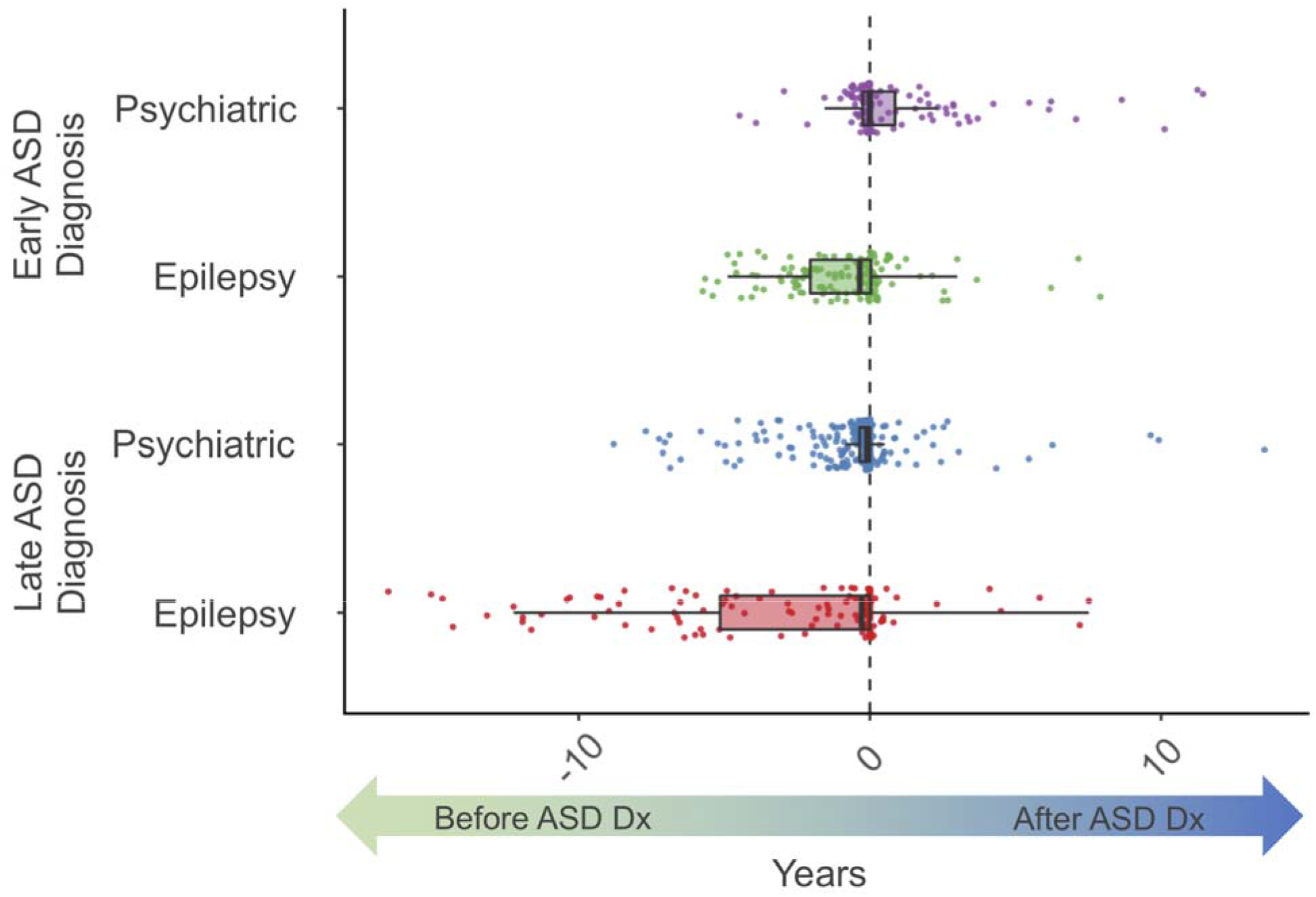
Time between first ASD diagnostic code and first co-occurring condition code, for epilepsy or psychiatric conditions. Dashed line denotes age at first ASD diagnosis. On x-axis, negative values denote years *before* ASD diagnosis, positive values denote years *after* ASD diagnosis.

We also investigated if the age of first diagnostic code in our dataset for epilepsy and psychiatric conditions differed in ASD compared to controls (**Figure 9**). The onset of epilepsy-related conditions was later in ASD compared to the TD group: early diagnosis group, 3.36±2.27 years compared to 2.23±2.35 years, W=7875, p=6.126e-06; late diagnosis group, 10.77±6.74 years compared to 8.55±9.95 years, W=4466, p=0.0017. The onset of psychiatric conditions was *earlier* for those with ASD compared to TD, but only in the early diagnosis group, 5.56±2.54 years compared to 7.48±3.74 years, W=1721, p=0.0008. In the late ASD group there was no difference in onset of psychiatric conditions, 13.02±6.77 years compared to 15.76±10.66 years, W=12534, p=0.1157.

**Figure 9.**
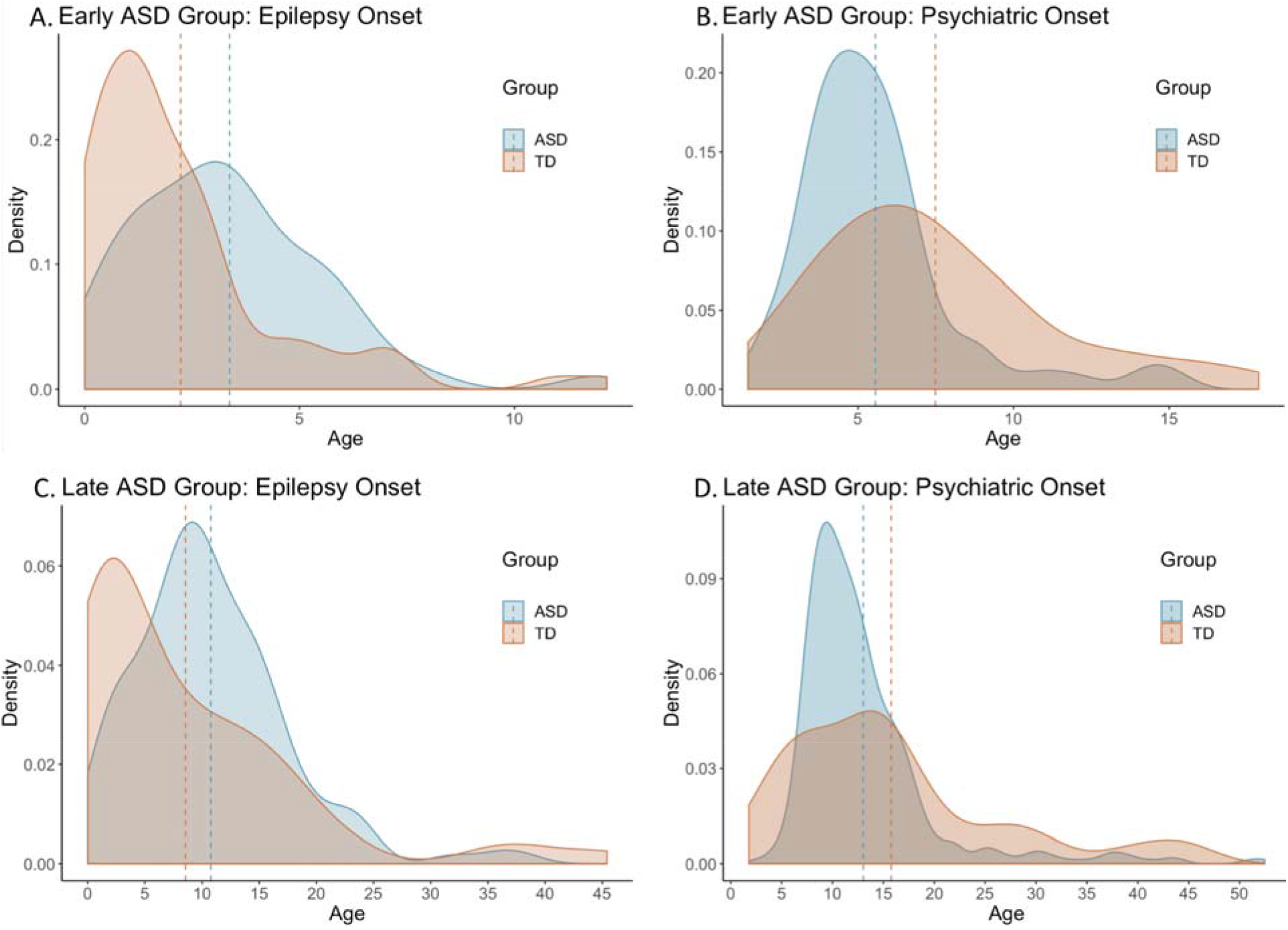
Onset of epilepsy-related (A,C) or psychiatric (B,D) conditions within ASD cohorts (early, A,B; late, C,D) compared to matched controls. Dashed lines indicate mean onset for each group. Onset, or first code in our dataset, of epilepsy-related conditions is older for ASD compared to matched controls: early, 3.36±2.27 compared to 2.23±2.35, W=7875, p=6.126e-06; late, 10.77±6.74 compared to 8.55±9.95, W=4466, p=0.0017. Onset of psychiatric conditions was younger for ASD compared to matched controls in the early group, 5.56±2.54 compared to 7.48±3.74, W=1721, p=0.0008; but there was no group difference in the late ASD group, 13.02±6.77 compared to 15.76±10.66, W=12534, p=0.1157.

## Discussion

This study characterizes co-occurring medical conditions in ASD with a new tool (pyPheWAS), allowing us to understand the burden of these conditions in ASD through the presence, count, and duration of related visits. This work provides strong evidence of an increased medical burden on individuals with ASD and highlights the need for adequate management of co-occurring conditions. By characterizing this burden, especially by age at ASD diagnosis, we can build predictive models of co-occurring condition progression (eg. onset peaks), introduce targeted risk assessments across the lifespan, and move towards more personalized medicine, ultimately reducing burden.

This study confirms many previously reported co-occurring condition categories in ASD, including neurological^5,6,18^, psychiatric^8,18^, and developmental conditions^6,18^. A particularly burdensome co-occurring condition for individuals and their families is disordered sleep^29,30^, with reported rates of 50-80%^31^ in ASD. Sleep disturbances exacerbate core symptoms of ASD^32^, which could explain increased visits and longer durations associated with sleep disorders in our study. Epilepsy and related conditions were also elevated in ASD. The prevalence of epilepsy in ASD (5-38%^5,33^) is significantly higher than the general population (1.2%^34^). There may be two epilepsy-onset peaks in ASD, before age 5, and in adolescence^35^. In our study, these peaks were present, and differed from the control group. Underlying biological phenomena may explain overlapping diagnostic peaks in epilepsy and ASD^36^. Yet, co-occurring diagnoses may also reflect increased medical care usage, increasing the likelihood of identification for ASD. In our analysis, only *prevalence* of epilepsy was increased in ASD, suggesting that while individuals with ASD are more likely to have epilepsy, the management and progression of epilepsy-related conditions may not differ in ASD.

We observed significant co-occurrence of psychiatric conditions in ASD, consistent with previous reports^8,37,38^. Co-occurring psychiatric conditions represent a significant burden for individuals with ASD and their families as they are related to emergency department visits^16,39^, associated with parental stress^40^, and remain prevalent throughout adulthood^41^. Psychiatric disorders were also associated with more visits and longer durations in ASD. Some conditions, such as ADHD, were only associated with ASD when examining duration of co-occurring conditions. There may be multiple explanations for this phenomenon: psychiatric conditions may present earlier or result in more severe complications in ASD. Alternatively, this could reflect closer monitoring for other psychiatric conditions after an ASD diagnosis.

Our findings of increased presence of depression and anxiety disorder in ASD are consistent with smaller studies of anxiety (11-84%^42,43^) and depression (1-76%^10,44^). We also observed co-occurring suicidal ideation in ASD. Recent studies report elevated levels of suicidal ideation in ASD^45–47^ and the field has now begun to recognize this issue^48^. Adjustment disorders are associated with suicidal behaviors in ASD^49,50^, but the contribution of anxiety and depression is not known. Given elevated rates of depression in ASD^44^, it will be imperative to understand mood disorders and suicidal ideation in the context of ASD *explicitly*, allowing for identification of unique risk factors and tailored treatments.

We also examined differences in co-occurring medical conditions by age at ASD diagnosis. Estimates of average age of ASD diagnosis^19^ suggest a peak around 3-4 years old^51^, when stable diagnoses can be made, but as many as half of cases may not be identified until age six or later, and a significant number of individuals are not diagnosed until even later childhood or adulthood^20,21,24^. Many hypothesize these diagnostic peaks reflect ASD subtypes, while others suggest a “two-hit” hypothesis, where adolescence, with its increased social demands and biological changes, may overwhelm adaptive functioning^52^. The “two-hit” hypothesis could explain why we observed more girls/women in the later diagnostic group; women may be able to camouflage ASD symptoms until adolescence^53^. Consistent with our data, women may also be more likely to receive other psychiatric diagnoses prior to their ASD diagnosis^54^.

In our analysis, we describe co-occurring condition profiles by age at diagnosis that diverge around epilepsy-related and psychiatric conditions. Similar subtypes have been reported in ASD^17^, with one study reporting both a psychiatric and an epilepsy-enriched subtype^11^, among other subtypes. In another study, co-occurring psychiatric conditions were present in 77% of individuals with higher cognitive functioning and later ages of ASD diagnosis (∼11 years), with up to ∼20% of co-occurring conditions potentially mis-diagnosed^38^. It is not clear if these differences by diagnostic group reflect subtypes with different underlying etiologies or if later diagnoses represent more complicated cases. Later ASD diagnosis could be due to other factors (like socioeconomic status) that we could not test using EMR, but may contribute to misdiagnosis or delayed evaluation^55^. Similarly, we observed different onset patterns for epilepsy and psychiatric conditions, compared to controls and by age at ASD diagnosis. More work is needed to determine the causes of altered diagnostic timing in ASD-related co-occurring conditions.

We also identified phecodes that were *less* likely to be associated with ASD (see **Table 2**). These findings could suggest individuals with ASD are less likely to experience sore throats, injuries, and acute pain; yet, evidence suggests altered pain responsivity, reporting, and expression in ASD^56,57^, possibly reducing identification of conditions that require pain reporting. Motivation to seek medical care may also be reduced in individuals with ASD as medical facilities can exacerbate sensory sensitivities and increase social demands, complicating assessment^1^. Alternatively, these conditions that are less present in the ASD group could reflect potential catchment effects of the control group.

There are some limitations to consider in this study. As ICD-9 codes are primarily used for billing purposes, they do not represent a definitive diagnosis^58^. However, our use of phecodes, which group relevant ICD-9 codes, increases our diagnostic confidence in each condition. Similarly, these data may not represent the larger population and may reflect some catchment artifacts. We have reduced the likelihood of these artifacts with rigorous control matching. This study also has a number of strengths. The PheDAS tool allowed for a nuanced characterization of co-occurring medical conditions across the lifespan of individuals with ASD, beyond simply prevalence of each condition. The ability to examine count and duration of conditions allows us to better understand the burden of these conditions in ASD. This study confirms previous reports^2,4,11^, extending our understanding of medical issues in ASD, but more importantly, highlights the urgency to reduce the co-occurring condition burden for people with ASD.

## Data Availability

The datasets used for the analyses described were obtained from Vanderbilt University Medical Center?s BioVU and are not available for distribution.

## Acknowledgements

The datasets used for the analyses described were obtained from Vanderbilt University Medical Center’s BioVU which is supported by numerous sources: institutional funding, private agencies, and federal grants. These include the NIH funded Shared Instrumentation Grant S10RR025141; and CTSA grants UL1TR002243, UL1TR000445, and UL1RR024975. Dr. Failla was supported by a NIMH training grant (T32-MH18921) during completion of this work. This work was also supported in part by the National Institute of Biomedical Imaging and Bioengineering training grant T32-EB021937 and by Eunice Kennedy Shriver National Institute of Child Health & Human Development grant U54HD08321105.

## Notes

### Competing Interest Statement

The authors have declared no competing interest.

### Author Declarations

All relevant ethical guidelines have been followed and any necessary IRB and/or ethics committee approvals have been obtained.

Any clinical trials involved have been registered with an ICMJE-approved registry such as ClinicalTrials.gov and the trial ID is included in the manuscript.

